# Virologic outcomes on dolutegravir-, atazanavir-, or efavirenz-based ART in urban Zimbabwe: A longitudinal study

**DOI:** 10.1101/2023.10.08.23296721

**Authors:** Tinei Shamu, Matthias Egger, Tinashe Mudzviti, Cleophas Chimbetete, Justen Manasa, Nanina Anderegg

## Abstract

There are few data from sub-Saharan Africa on the virological outcomes associated with second-line ART based on protease inhibitors or dolutegravir (DTG). We compared viral load (VL) suppression among people living with HIV (PLWH) on atazanavir (ATV/r)- or dolutegravir (DTG)-based second-line ART with PLWH on efavirenz (EFV)-based first-line ART. We analyzed data from the electronic medical records system of Newlands Clinic in Harare, Zimbabwe. We included patients aged ≥12 years when commencing first-line EFV-based ART or switching to second-line DTG- or ATV/r-based ART with ≥24 weeks follow-up after start or switch. We computed suppression rates (HIV VL <50 copies/mL) at weeks 12, 24, 48, 72, and 96 and estimated the probability of VL suppression by treatment regimen, time since start/switch of ART, sex, age, and CD4 cell count (at start/switch) using logistic regression in a Bayesian framework. We included 7013 VL measurements of 1049 PLWH (61% female) initiating first-line ART and 1114 patients (58% female) switching to second-line ART. Among those switching, 872 (78.3%) were switched to ATV/r and 242 (21.7%) to DTG. VL suppression was lower in second-line ART patients than first-line ART patients, except at week 12, when those on DTG showed higher suppression than those on EFV (aOR 2.10, 95%-credible interval [CrI] 1.48-3.00) and ATV/r-based regimens (aOR 1.87, 95%-CrI 1.32-2.71). In weeks ≥48, first-line patients had around 3 times the odds of VL suppression compared to second-line patients. There was no evidence of a difference between DTG and ATV/r for follow-up times ≥24 weeks in PLWH on second-line. Second-line ART patients had lower VL suppression rates than those receiving first-line ART, and there was no difference in suppression between DTG- and ATV/r-based second-line ART ≥6 months after switching. Virologic monitoring and adherence support remain essential to prevent second-line treatment failure in settings with limited treatment options.

## Introduction

Antiretroviral therapy (ART) has become more effective, more affordable, and safer since the advent of combination therapy in the mid-90s [1]. Currently, the integrase inhibitor dolutegravir (DTG) in combination with two nucleoside reverse transcriptase inhibitors (NRTIs) is recommended for patients initiating ART and for second-line ART among patients who experienced virologic failure on regimens that did not include DTG [2]. Previously, efavirenz (EFV) was recommended for first-line, and the ritonavir-boosted protease inhibitors atazanavir (ATV/r) or lopinavir (LPV/r) for second-line ART, always combined with two NRTIs. DTG achieves earlier virologic suppression compared to ritonavir-boosted protease inhibitors, has fewer toxicities and may have a lower or similar incidence of resistance among patients receiving second-line ART [2–4]. Compared with efavirenz, DTG had superior viral suppression over 96 weeks, was protective against drug resistance, and led to fewer discontinuations [5–7].

The success of second-line ART depends on several factors. Poor adherence to treatment has been identified in many studies as the critical determinant of virologic failure and the emergence of drug resistance [8,9]. A high viral load, a low CD4 cell count or advanced HIV infection at the time of switching, concomitant treatment for tuberculosis, and younger age are also associated with poorer outcomes on second-line ART [10–13]. The availability of DTG in a fixed dose combination with lamivudine (3TC) and tenofovir disoproxil fumarate (TDF), commonly referred to as TLD, may improve treatment compliance and lead to better virologic suppression rates among people living with HIV (PLWH) on second-line ART [2].

Compared with boosted protease inhibitors for ART naïve and treatment-experienced PLWH with background NRTI resistance, DTG was neither inferior nor superior for viral suppression [14–16]. In PLWH switching to DTG-based second-line ART, viral replication is typically reduced to undetectable levels within the first four to 24 weeks of therapy [17,18]. However, long-term real-world virologic suppression and clinical outcomes after switching to second-line ART are less well documented. Indeed, there is a lack of data on the outcomes associated with second-line ART based on boosted protease inhibitors and second-line and first-line ART based on DTG-containing regimens, particularly in sub-Saharan Africa. We compared virologic suppression among adolescents and adults switching to DTG- or ATV/r-based second-line ART after experiencing virologic failure with those starting EFV-based first-line ART in an HIV care and treatment program in Zimbabwe.

## Methods

### Setting and data

We analyzed data of patients receiving ART at Newlands Clinic in Harare, Zimbabwe. Newlands Clinic is an outpatient HIV referral centre with approximately 7,300 patients in care as of June 2022. The clinic provides comprehensive HIV care to clients of low socio-economic status, including ART, laboratory monitoring, psychosocial support, reproductive health care and other ancillary services. It is supported by the Ruedi Luethy Foundation, a Swiss-based private voluntary organization, and part of the International epidemiology Databases to Evaluate AIDS (IeDEA) in Southern Africa. More details on the clinic’s activities are provided elsewhere [19,20]. From 2010 to 2019, EFV-based ART was predominantly used as first-line ART and ATV/r as second-line ART at Newlands Clinic. Since 2019, DTG-based ART has been used both in first- and second-line ART [2,21].

The present analysis was based on the database compiled from the clinic’s electronic medical records system. The system is password protected, and only approved clinic staff can log in and view patient records in accordance with the level of permission. We abstracted longitudinal data of patients receiving ART between February 2013 and June 2022, including demographic information, CD4 cell counts, and HIV-1 RNA viral loads. Viral loads were typically measured at the time of switching or starting (baseline). After baseline, viral loads were measured at weeks 12 and 24 and then every 24 weeks. Measurements were done using the COBAS Ampliprep/Taqman48 platform and Roche HIV-1 version 1.0 kits. CD4 counts were measured at baseline using a Partec Cyflow Counter II machine with CD4 Easy Count reagents by Sysmex.

Patients enrolling into care at Newlands Clinic provided informed written consent that allows the use of data accumulating during their routine care for research under the IeDEA collaboration. IeDEA was approved by the Medical Research Council of Zimbabwe (MRCZ No. A1336) [22]. The study data were stored in a password protected electronic medical records system and were accessed on the 30^th^ of August 2022 by authorised clinic personnel. The abstracted data were supplied to the statistical analysis team as de-identified records that could not be traced to individual patients.

### Inclusion criteria

We included patients aged 12 years and older at the time of commencing ART or switching to second-line ART who had at least 24 weeks on the new ART regimen. For patients commencing ART, we included those starting with a regimen containing EFV, lamivudine (3TC) and one of tenofovir (TDF), zidovudine (AZT), or abacavir (ABC). For second-line ART, we included patients switching with a viral load above 200 copies/mL from an NNRTI-based first-line to second-line regimens containing DTG or ATV/r combined with an NRTI backbone, as described above.

### Outcome

The outcome of interest was viral load suppression (HIV viral load <50 copies/mL) at week 12, 24, 48, 72 and 96 after baseline, in line with the clinic visits with scheduled VL measurements. We allowed for a one-month window around these time points to account for shifts in visit dates. For example, the “Week 24” measurements included VL measurements done 24 ± 4 weeks after baseline. If several measurements were available, the one closest to the visit date was chosen for analysis. Some VL measurements fell between two windows (for example, a measurement drawn 35 weeks after baseline was neither assigned to “Week 24” nor to “Week 48”). These measurements were used to impute missing VL information when a patient did not have a VL measurement for a visit date but did have a measurement right before and after the window around the visit date. We assumed that the missing measurement indicated suppressed or unsuppressed VL if both measurements (before and after) showed a suppressed or unsuppressed VL. If the measurements before and after were different (suppressed and unsuppressed), we did not impute the VL suppression information for the visit date.

### Explanatory variables

Explanatory variables included the time of the VL measurement (as defined above as Week 12, Week 24, Week 48, Week 72, or Week 96 after baseline), the ART regimen, the patient’s sex, age and CD4 cell count at baseline. Age at baseline was grouped into 12-19 years, 20-29, 30-39 years and ≥40 years. CD4 cell count at baseline was grouped into 0-199 cells/mm^3^, 200-349 cells/mm^3^ and ≥350 cells/mm^3^. ART regimens included “EFV” for first-line ART, and “ATV/r” and “DTG” for second-line ART.

### Statistical analysis

We calculated crude proportions of viral load suppression after baseline by ART regimen. We then estimated the probability of viral load suppression using multivariable logistic regression models in a Bayesian framework. To account for the dependence of viral load measurements within patients, we included a random intercept by patient. As fixed effects, we included the covariates ART regimen, time since baseline, sex, age and CD4 cell count. We checked if the inclusion of any two-way interaction between covariates improved the model fit in two steps. First, we compared model fits of the simple model without any interactions to alternative models containing one single two-way interaction. All two-way interactions for whom the comparison to the simple model showed a difference in expected log pointwise density that was larger two times its standard error were considered “eligible” for inclusion in the final model [23]. In the second step, we looked at all possible combinations of the eligible two-way interactions. The final model was the one with the best fit (again, according to the expected density).

We report adjusted odds ratios (aOR) for VL suppression and crude and predicted proportions of VL suppression with 95% credible intervals (CrIs). For both aORs and predicted probabilities, we report marginalized (“sample-averaged”) estimates derived by integrating over the distribution of random effects. Next to reporting predicted proportions stratified by all levels of covariates, we also report them standardized and non-standardized by time and treatment regimen. For standardized predictions, we predicted the probability of VL suppression at weeks 12, 24, 48, 72, and 96 for every study patient, assuming they were on DTG, then assuming they were on ATV/r and finally on EFV. We then averaged all the predicted probabilities of viral load suppression for each treatment regimen and time point. Non-standardized predictions consisted of predicting the probability of VL suppression for the observed data only.

## Results

Overall, we included 2163 PLWH (59.9% female), of whom 1114 were switching to second-line ART, and 1049 were initiating first-line ART (Table 1). Patients switching to second-line ART were younger (median age 29 years; IQR 19-42) than those starting first-line ART (36 years; IQR 28-44). Among the second-line ART patients, 872 (78.3%) switched to ATV/r-based ART, while 242 (21.7%) switched to DTG-based ART (Table 1). Patients switching to ATV/r-based second-line ART had lower CD4 cell counts than those switching to DTG or starting an EFV-based first-line regimen (Table 1). The 2163 patients contributed 7413 VL measurements, of which (7181, 96.9%) were observed and 232 (3.0%) imputed. Around three-quarters of patients had a VL measurement at shorter follow-up times (weeks 12, 24, 48), while this proportion was lower for longer follow-up times (Table 1). For DTG, fewer patients had a VL measurement at longer follow-up times than the other two regimens. As indicated by the later calendar years of starting/switching ART for DTG, this was mainly due to the follow-up time of patients switching to DTG being too short to contribute VL measurements for later weeks (Table 1). Only a few patients were lost to follow-up or died during the follow-up time relevant to this study (Table 1).

**Table 1.** Baseline demographic and clinical characteristics of patients by treatment regimen.

|  | First line | Second line |  | Overall |
| --- | --- | --- | --- | --- |
|  | EFV | ATV/r | DTG |  |
| **Total** | 1049 | 872 | 242 | 2163 |
| **Female** | 645 (61.5%) | 506 (58.0%) | 145 (59.9%) | 1296 (59.9%) |
| **Male** | 404 (38.5%) | 366 (42.0%) | 97 (40.1%) | 867 (40.1%) |
| **Age [years]** |  |  |  |  |
| **Median (IQR)** | 36 (28-44) | 29 (19-41.25) | 28.5 (19-42) | 34 (22-43) |
| **12 - 19** | 97 (9.2%) | 224 (25.7%) | 67 (27.7%) | 388 (17.9%) |
| **20 - 29** | 202 (19.3%) | 219 (25.1%) | 58 (24.0%) | 479 (22.1%) |
| **30 - 39** | 348 (33.2%) | 161 (18.5%) | 49 (20.2%) | 558 (25.8%) |
| **40+** | 402 (38.3%) | 268 (30.7%) | 68 (28.1%) | 738 (34.1%) |
| **CD4 cell count [cells/mm<sup>3</sup>]** |  |  |  |  |
| **Median (IQR)** | 251 (122-363) | 182 (73-331) | 253 (78-469) | 224 (97-360) |
| **<200** | 426 (40.6%) | 461 (52.9%) | 98 (40.5%) | 985 (45.5%) |
| **200-349** | 331 (31.6%) | 219 (25.1%) | 51 (21.1%) | 601 (27.8%) |
| **>=350** | 292 (27.8%) | 192 (22.0%) | 93 (38.4%) | 577 (26.7%) |
| **Number of viral load measurements** |  |  |  |  |
| **Median (IQR)** | 4 (2-4) | 4 (3-5) | 3 (2-4) | 4 (3-4) |
| **Viral load suppression** |  |  |  |  |
| **at week 12** | 800 (76.3%) | 683 (78.3%) | 202 (83.5%) | 1685 (77.9%) |
| **at week 24** | 884 (84.3%) | 722 (82.8%) | 175 (72.3%) | 1781 (82.3%) |
| **at week 48** | 812 (77.4%) | 663 (76.0%) | 155 (64.0%) | 1630 (75.4%) |
| **at week 72** | 424 (40.4%) | 506 (58.0%) | 108 (44.6%) | 1038 (48.0%) |
| **at week 96** | 627 (59.8%) | 565 (64.8%) | 87 (36.0%) | 1279 (59.1%) |
| **Year of ART start/switch** |  |  |  |  |
| **Median** | 2016 | 2017 | 2021 | 2017 |
| **(IQR)** | (2015-2017) | (2016-2018) | (2020-2021) | (2016-2018) |
| **Status at end of follow-up\*** |  |  |  |  |
| **In care** | 988 (94.2%) | 832 (95.4%) | 230 (95.0%) | 2050 (94.8%) |
| **LTFU** | 42 (4.0%) | 18 (2.1%) | 4 (1.7%) | 64 (3.0%) |
| **Dead** | 19 (1.8%) | 22 (2.5%) | 8 (3.3%) | 49 (2.3%) |

\* LTFU and Dead correspond to patients who were lost to follow-up or died within the follow-up time relevant to this study. In care corresponds to those who remained in care by week 100 after starting/switching ART or at database closure, whichever came first. EFV, efavirenz; ATV/r, ritonavir-boosted atazanavir; DTG, dolutegravir; IQR, interquartile range; LTFU, lost to follow-up.

Crude rates of virologic suppression were lower on second-line ART than first-line ART (Figure 1). The exception was week 12 when the DTG-based regimen showed higher suppression (crude proportion 69.9%, 95%-CrI 63.6-75.9%) than ATV/r (crude proportion 53.9%, 95%-CrI 50.1-57.8%) and EFV-based regimens (crude proportion 57.1%, 95%-CrI 53.6-60.5%). Among the second-line ART regimens, crude rates of virologic suppression for follow-up times greater than 24 weeks were slightly higher for DTG (73.5%, 95%-CrI 69.7-77.5%) compared to ATV/r (68.2%, 95%-CrI 66.2-70.0%).

**Figure 1.**
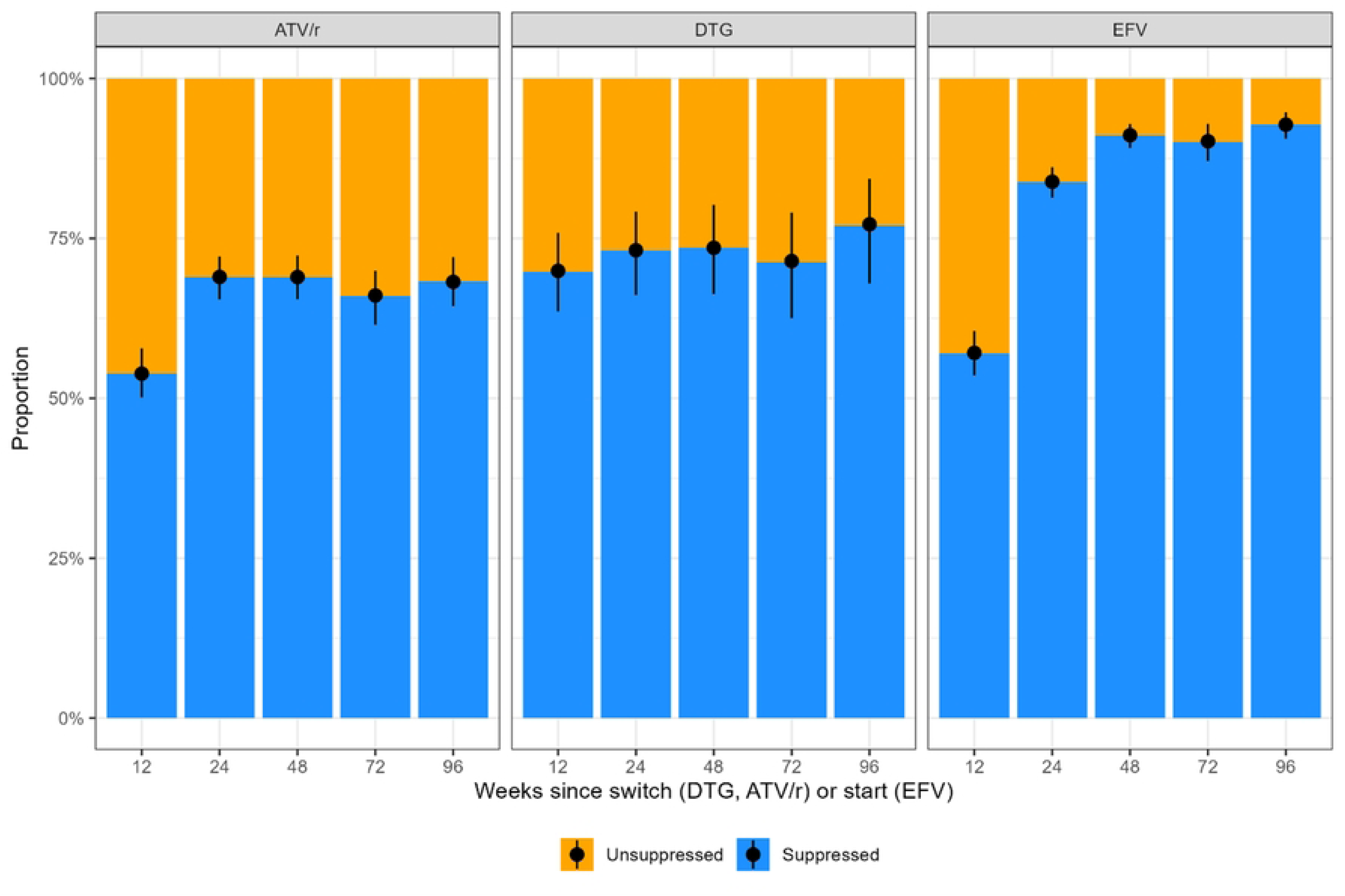
Crude proportions (with 95% credible intervals) of viral load suppression by treatment regimen and time post switch (ATV/r, DTG) or start (EFV) of ART. EFV, efavirenz; ATV/r, ritonavir-boosted atazanavir; DTG, dolutegravir.

Only the two-way interactions between time and ART regimen and between time and age met the criteria for potential inclusion in the final model. The model including only the time-ART regimen interaction, and the model including both the time-ART regimen and the time-age interaction led to similar model fits, with a slightly better fit for the simpler model (Supplementary Table S1). The final logistic regression model thus only included the two-way interaction between time and ART regimen. The final model fitted the data well, with a few exceptions when there was large uncertainty due to small numbers (Supplementary Figure S1).

The regression model confirmed the higher odds of VL suppression for DTG at short follow-up (aOR 1.87, 95% CrI 1.32-2.71 comparing DTG to ATV/r at Week 12 and aOR 2.10, 95% CrI 1.48-3.00 comparing DTG to EFV, Figure 2). While for DTG, the odds of VL suppression did not increase substantially for longer follow-up periods, they did so for patients on ATV/r and even more for those on EFV (Figure 2, Figure 3). After 24 weeks, the estimated odds of VL suppression were relatively stable for both second-line regimens, and there was no evidence of a difference between DTG and ATV/r. For EFV, the odds of VL suppression also reached a stable plateau but slightly later, at 48 weeks. At that time, the odds of VL suppression was estimated to be around 3 times higher than the second-line regimens (Figure 2, Figure 3). Apart from the differences between first- and second-line regimens, the odds of VL suppression were higher for females and increased with age and CD4 cell count (Figure 2). VL suppression was lowest for adolescents, with estimated suppression rates mostly below 75% in adolescents switching to second-line ART and below 60% for those switching with CD4 cell counts below 200 cells/mm^3^ (Supplementary Figure S1).

**Figure 2.**
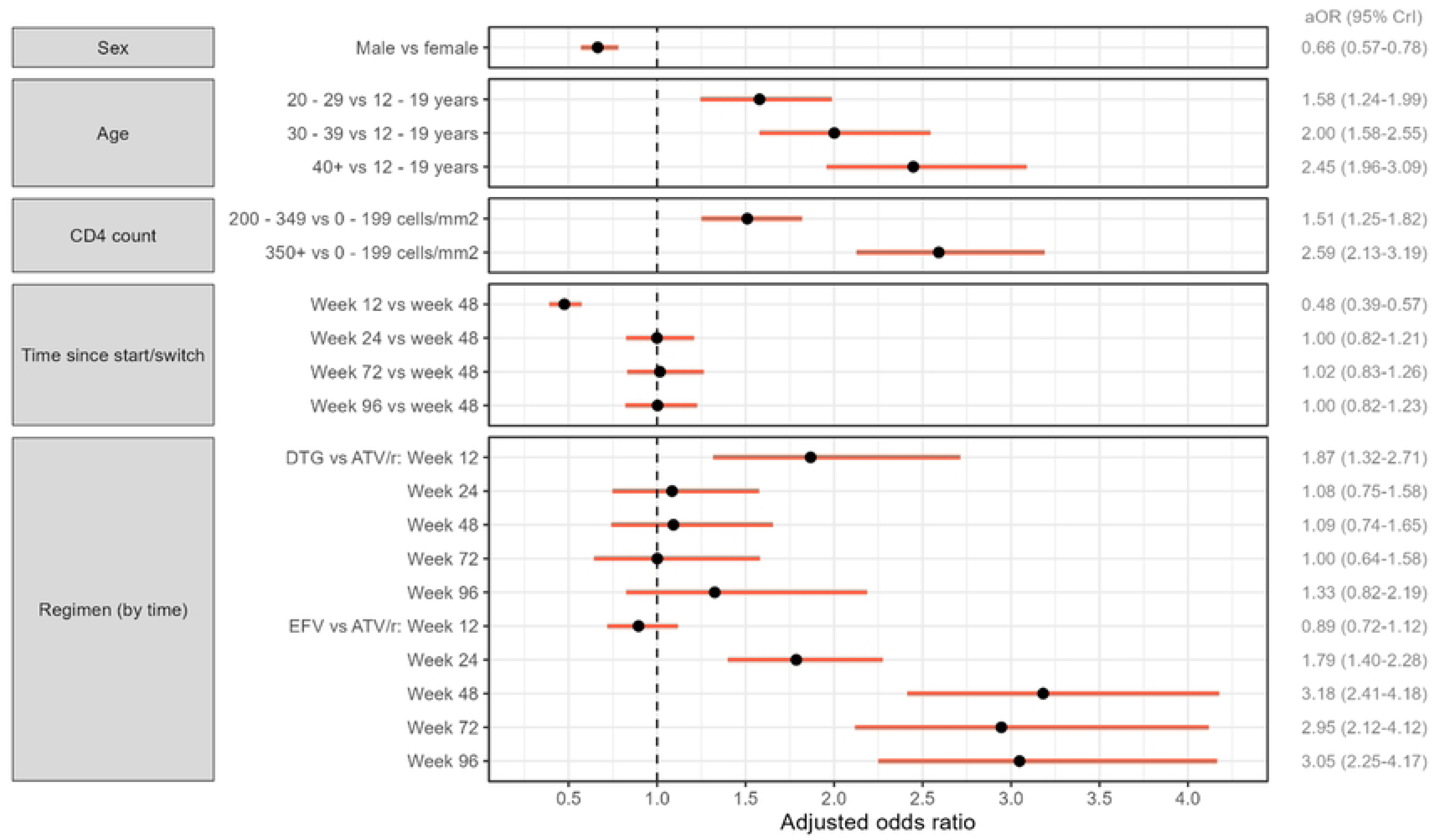
Adjusted odds ratios (aOR) for viral load suppression with 95% (red) credible intervals. Results derived from a Bayesian logistic regression model including a random intercept by patient and the covariates sex, baseline age, baseline CD4 cell count, time since start/switch, and ART regimen. The model also includes a two-way interaction between ART regimen and time since start/switch, so that the aORs comparing ART regimens are shown stratified by time. Estimates are marginalized by integration over the distribution of random effects. EFV, efavirenz; ATV/r, ritonavir-boosted atazanavir; DTG, dolutegravir.

**Figure 3.**
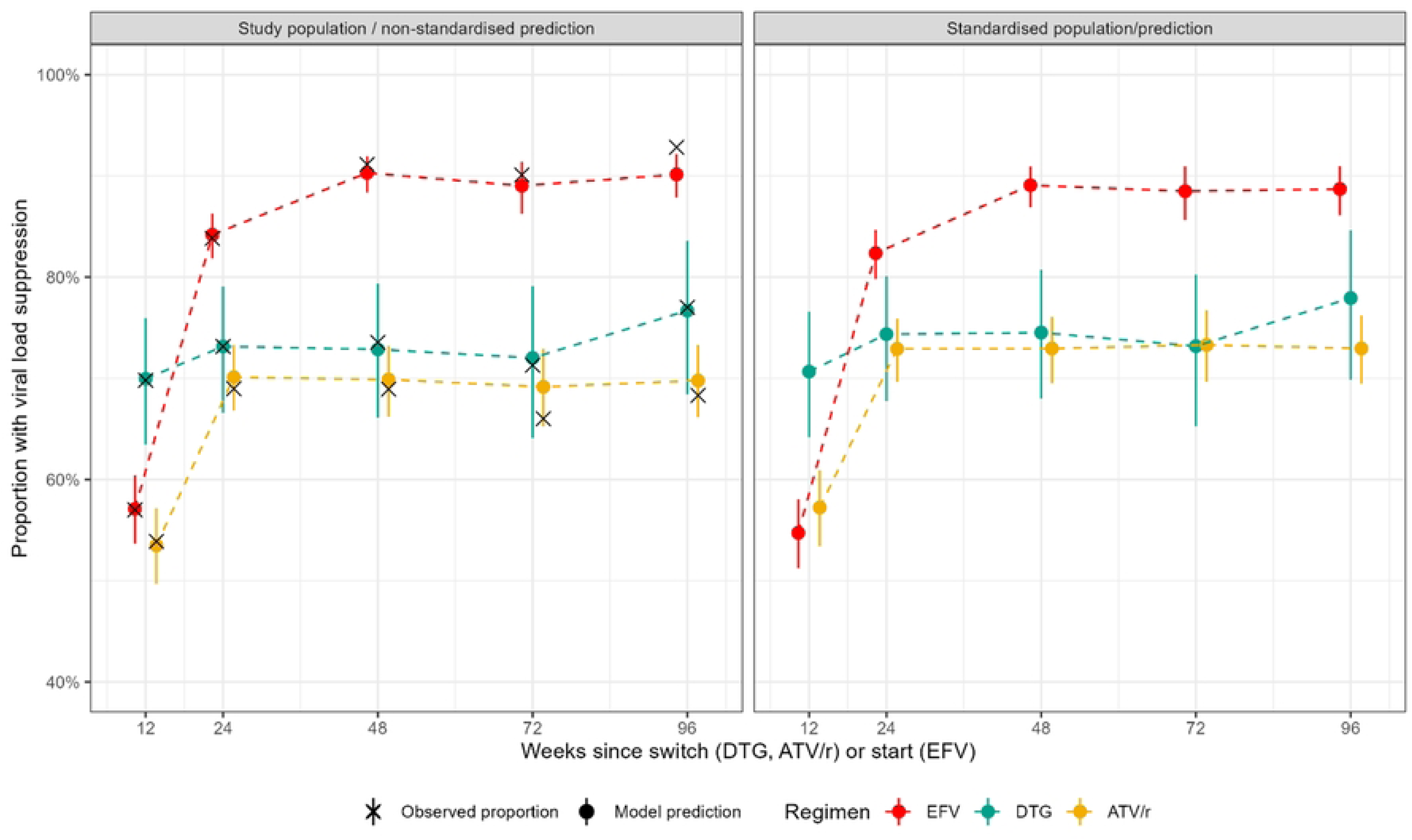
Non-standardized (left panel) and standardized (right panel) predicted proportions and 95% credible intervals of viral load suppression by treatment regimen and time since switch (DTG, ATV/r) or start (EFV). Results from a Bayesian logistic regression model including a random intercept by patient and the covariates sex, baseline age, baseline CD4 cell count, time since start/switch, ART regimen, and a two-way interaction between time since start/switch and ART regimen. Estimates are marginalized by integrating over the distribution of random effects.

Standardized and non-standardized differences in VL suppression rates between regimens were overall similar, but standardization attenuated differences between the two second-line regimens (Figure 3). At week 48, standardized predicted proportions of VL suppression were 89.1% (95% CrI 86.9-90.9%) for EFV, 74.5% (95%-CrI 68.0-80.7%) for DTG, and 72.9% (95%-CrI 69.5-76.1%) for ATV/r (Figure 3). Corresponding non-standardized proportions of VL suppression at week 48 were 90.3% (95%-CrI 88.4-91.9%) for EFV, 72.9% (95%-CrI 66.1-79.3%) for DTG, and 69.9% (95%-CrI 66.2-73.2%) for ATV/r (Figure 3).

## Discussion

In this longitudinal analysis of routine clinical data collected in an urban ART program in Zimbabwe, we examined virologic suppression among PLWH switching to DTG- or ATV/r-based second-line ART after experiencing virologic failure, and PLWH starting ART on EFV-based first-line regimens. Suppression rates were highest for first-line patients receiving EFV-based ART from weeks 24 through 96. Older age and higher CD4 cell count at switch or start were associated with higher odds of virologic suppression. In contrast, male sex was associated with reduced odds of virologic suppression across all regimens. Our results confirm the more rapid suppression of viral replication with DTG-based ART than EFV-based ART, even if used in second-line ART [3,4].

Our study’s limitations include the absence of objective measures of treatment adherence and baseline drug resistance profiles. Also, not all patients had VL measurements taken at all time points, especially in later weeks and patients switching to DTG. This was mainly due to the shorter follow-up time of patients switching to DTG. Contrary to EFV and ATV/r, which were mainly used before 2019 and thus allowed most patients to have ≥2 years of follow-up, DTG was only introduced in 2019. Therefore, recently switched patients can only contribute VL measurements to the earlier weeks. Further, many patients switched to DTG-based second-line during the Covid-19 national lockdown period. The different periods are a limitation of this study, as we cannot exclude residual confounding by the Covid-19 pandemic or other factors that might have changed over time.

Our study took advantage of a long-standing, well-characterized cohort with high retention in care and reliable data collection [20]. This is reflected in the fact that we had little loss to follow-up over the study period and very few missing CD4 counts, which is rare for observational cohorts in similar settings. Another strength lies in using a Bayesian approach, which allowed us to test for many interactions and calculate standardized probabilities of viral load suppression that allow more informative comparisons between treatment regimens correcting for differences in characteristics between first- and second-line patients.

Many of the PLWH on second-line ART will probably have a history of poor adherence, which led to virologic failure on first-line ART and, in some patients, could have led to the development of drug resistance. However, NNRTI pretreatment resistance likely played a more significant role in the last decade [24–27]. Second-line virologic failure (DTG or ATV/r) may more likely be related to poor treatment compliance based on literature showing almost no pretreatment or transmitted drug resistance among protease inhibitor- and INSTI-naïve PLWH in Zimbabwe and the demonstrated efficacy of both DTG and protease inhibitors with suboptimal NRTI backbones [15,25,28–30]. In addition, a longer duration of ART is associated with a higher likelihood of treatment failure [31,32]. Even though both DTG and ATV/r are potent antiretrovirals, with DTG being superior to EFV for both viral suppression and tolerability [5,7,33], the overall suppression rates among the second-line patients were lower than those observed for first-line patients in all age groups, sex, and baseline CD4 cell count strata. This suggests that for patients with a history of treatment failure, switching to DTG or ATV/r alone without other adherence support interventions may not be sufficient to reach the programmatic targets of 95% or higher VL suppression [34,35]. Failure to achieve virologic suppression among patients receiving second-line DTG-based ART can expose these patients to the emergence of drug resistance against DTG and the beginning of transmitted drug resistance to the INSTI class. Several cases of acquired and pretreatment DTG resistance have already been reported [36–38].

Almost half of the patients analyzed in this study had advanced HIV disease with baseline CD4 cell counts <200cells/mm^3^. Although lower CD4 cell counts are the consequence of HIV replication, it is unclear why lower baseline CD4 counts are associated with failure to suppress VL across all the regimens in this study. Higher viral loads at ART commencement are associated with prolonged durations before viral suppression leading to increased odds of virologic failure on NNRTI containing ART [39,40]. Further, severe immune suppression might indicate a lower commitment to starting/switching ART or engaging in adherence support initiatives, which in turn is likely associated with reduced treatment adherence [35]. Low CD4 cell counts when switching to second-line ART may also result from previous loss to follow-up, treatment interruption, or inconsistent clinic visits. Some studies have also shown a relationship between low baseline CD4 cell counts and subsequent virologic failure, while another study from Mozambique showed an association between higher baseline CD4 cell counts and virologic failure [24,41,42].

Patients switching to second-line ART were younger than those starting first-line. Likewise, younger patients were also less likely to achieve virologic suppression across all regimens. Younger age has previously been associated with poorer adherence and virologic failure without accompanying drug resistance [24,43]. This observation warrants investment in custom treatment support interventions that foster improved treatment adherence among young PLWH.

## Conclusion

Patients receiving second-line ART were less likely to achieve virologic suppression than those on first-line ART, despite switching to robust or even superior regimens. The probability of virologic suppression increased with increasing age and baseline CD4 cell count. Further efforts are needed to enhance treatment adherence among young PLWH and, more broadly, those switching after virologic failure in settings with limited treatment options.

## Data Availability

All datasets are available from figshare.com

## Acknowledgements

We acknowledge Ruedi Luethy and the Newlands Clinic staff and clients for making this work possible.

## Supporting information

**S1 Table. Model comparison: Difference in expected log pointwise predictive density (ELPD) and its standard error for a new dataset estimated by approximate leave-one out cross validation.**

**S2 Figure. Predicted (colour) and crude/observed (black) proportions (with 95% credible intervals) of viral load suppression stratified by all covariates included in the model fit.**

